# Drivers of Depression in Chronic Post-Stroke Aphasia

**DOI:** 10.1101/2024.11.13.24317297

**Authors:** Devna Mathur, Sachi Paul, Andrew DeMarco, Peter Turkeltaub

## Abstract

**Background:** Approximately one third of stroke survivors develop post-stroke depression, which reduces quality of life. Previous studies have often focused on early phases of recovery and excluded people with significant aphasia. Here, we examine relationships of depression symptoms to demographic factors, and different types of stroke-related disability and impairment in adults with chronic left-hemisphere stroke and a history of aphasia.

**Methods:** 92 chronic left-hemisphere stroke survivors with a history of aphasia and 70 controls participated in this cross-sectional study. The Beck Depression Inventory-II (BDI-II) measured depression symptoms. The Stroke Impact Scale 3.0 (SIS) assessed stroke-related disability in Cognitive, Physical, and Social Participation domains, as well as self-perceived Recovery. The Western Aphasia Battery Aphasia Quotient and the NIH Stroke Scale total motor score measured language and motor impairment. Spearman correlations examined bivariate relationships between variables. Ordinal regression models examined group differences in BDI-II scores (Model 1), and disability and impairment factors that predicted BDI-II scores (Model 2), accounting for demographic factors and antidepressant medication status.

**Results:** BDI-II scores were on average 3.4 points higher in stroke survivors than controls. Model 1 confirmed that this difference was significant, and found that age was inversely related to BDI-II scores. Bivariate correlations demonstrated that higher BDI-II scores were related to lower SIS Cognitive, Social Participation, and Recovery scores. Model 2 found that these three measures independently predicted BDI-II scores.

**Conclusions:** The factors related to depression may differ depending on the nature of the stroke, the types of deficits experienced, and the phase of recovery. In the chronic phase of left hemisphere stroke with aphasia, cognitive and communication disabilities, social participation, and self-perceived recovery are the primary correlates of depression symptoms. These findings highlight the importance of assessing for depression even long after left hemisphere stroke, and suggest potential targets for psychotherapy to improve depression.

## Introduction

Approximately one third of stroke survivors experience post-stroke depression (PSD) (Hackett et al., 2005, 2014). PSD reduces quality of life (Villa et al., 2018) and is associated with increased risk of mortality and poorer overall recovery after stroke (Cai et al., 2019). It is crucial to examine predictors and risk factors of PSD in order to better understand its origins, which may aid in treatment. Recent literature on PSD has focused on epidemiology, possible treatments, and risk factors, but the factors that lead some stroke survivors to develop depression remain unclear.

Reviews have identified several predictors of PSD, including stroke severity, physical disability, cognitive impairment, history of psychiatric disorders, and lack of social support (Ayerbe et al., 2013; Ladwig et al., 2023). A study by Paolucci et al. (2006) observed that females were more at a risk for PSD compared to males, while other studies have found male gender to be a strong determinant of PSD (Srivastava et al., 2010). Similarly, a study by Alajbegovic et al. (2014) examining occurrence of depression after stroke found that younger age is associated with higher risk of PSD, whereas other studies have not found older age to be a significant predictor (Kutlubaev and Hackett, 2014; Towfighi et al., 2017). Clinical factors measured by stroke scales such as the National Institutes of Health Stroke Scale (NIHSS) have also been found to be related to PSD. Specifically, higher scores on the NIHSS, indicating more severe impairment, were found to be an independent predictor of PSD (Meng et al., 2017). Physical disability, social support, and communication impairments have also been identified as independent predictors of PSD in stroke samples from different settings (Ladwig et al., 2023). Some studies have also found acute cognitive impairment to be an important factor impacting depression symptoms after stroke (Nys et al., 2006). Although prior studies have identified various psychosocial and demographic predictors of PSD, studies have often focused on the acute/subacute phases of recovery. Relationships between demographic factors, stroke-related impairments, various types of disability, and depression symptoms in the chronic phase of stroke remain unclear.

One study examining the prevalence of post-stroke depression found that 70% of patients with aphasia meet the diagnosis for depression three months after their stroke (Kauhanen et al., 2000). Yet, critically, individuals with aphasia are often excluded from studies of PSD due to language comprehension concerns, so it is unclear if prior findings generalize to this population. A systematic review by Townend et al. (2007) found that around 71% of stroke depression studies excluded people with aphasia. This sampling bias leaves a major gap in the literature, as this exclusion limits the understanding of PSD in relation to treatment and diagnosis to only about one third of stroke survivors with aphasia (Flowers et al., 2016).

In this study, we examine relationships between demographic factors, different types of disability (i.e., functional limitations with particular activities, such as daily life communication) and impairment (i.e., a problem with body or mental function, such as measured aphasia severity), and symptoms of depression in adults with chronic left-hemisphere stroke and a history of aphasia. Previous studies of PSD have often used generic measures of health-related quality of life, such as the Short Form-36 Health Survey and the EuroQol Instrument, which is a general measure that is not intended to specifically assess the stroke population. We address this limitation by using the Stroke Impact Scale (SIS), a comprehensive measure of disability and health-related quality of life after stroke (Duncan et al., 1999), which few prior studies of PSD have used. We aim to better understand the possible demographic and clinical predictors of PSD symptoms in individuals with a history of post-stroke aphasia.

## Methods

### Participants

Participants were 92 individuals with left-hemisphere stroke and history of aphasia, all in the chronic phase (greater than 6 months), and 70 controls matched on age and education (See Table 1 for additional information about sample demographics). All participants were recruited for a study examining individual differences in language and cognition after left-hemisphere stroke (Clinicaltrials.gov NCT04991519). Stroke participants were considered eligible if they had a left-hemisphere stroke without clinically significant damage in other parts of the brain, and a history of aphasia either by self-report or based on medical records. Controls had no history of stroke. All participants were also required to meet the following criteria: over the age of 18, learned English at 8 years or younger, no history of significant neurological disease other than stroke (e.g., multiple sclerosis, premorbid mild cognitive impairment), no history of severe psychiatric conditions requiring hospitalization or ongoing use of psychiatric medications other than common antidepressants, and no history of learning disabilities requiring educational intervention (e.g., developmental dyslexia). Stroke survivors were required to score at least 4 out of 10 on the Western Aphasia Battery Auditory Verbal Comprehension subscore. Two individuals were not administered the Beck Depression Inventory-II (BDI-II) due to their low comprehension. From an initial sample of 95, one stroke survivor was excluded from analyses as they did not demonstrate evidence of aphasia based on assessment findings or medical records, and two stroke survivors were excluded due to stroke within the prior 6 months. All participants provided written informed consent as approved by the Georgetown University Institutional Review Board.

**Table 1.**
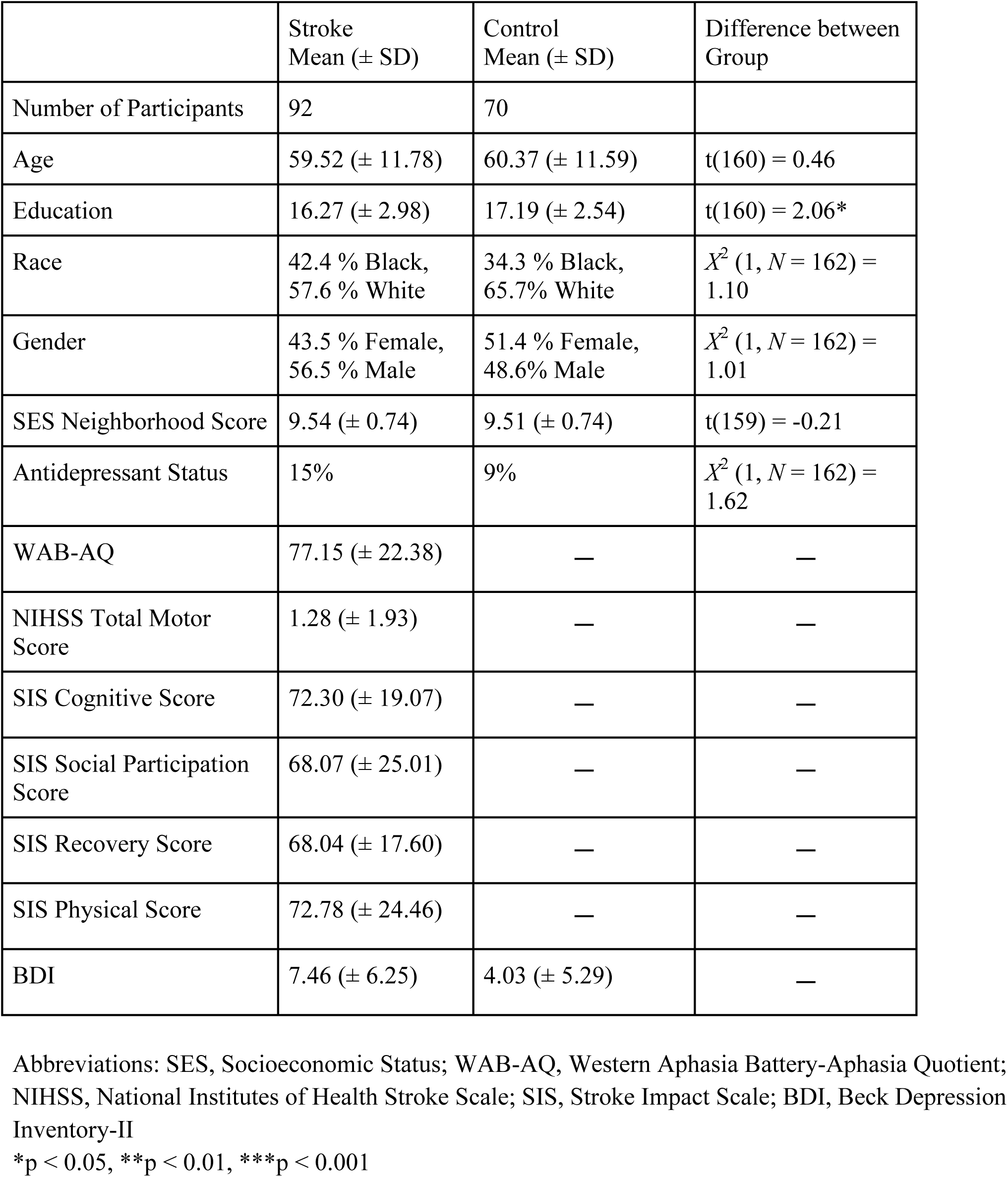
Sample characteristics.

| | Stroke<br>Mean (± SD) | Control<br>Mean (± SD) | Difference between<br>Group |
| --- | --- | --- | --- |
| Number of Participants | 92 | 70 |  |
| Age | 59.52 (± 11.78) | 60.37 (± 11.59) | $t(160) = 0.46$ |
| Education | 16.27 (± 2.98) | 17.19 (± 2.54) | $t(160) = 2.06^*$ |
| Race | 42.4 % Black,<br>57.6 % White | 34.3 % Black,<br>65.7% White | $X^2 (1, N = 162) = 1.10$ |
| Gender | 43.5 % Female,<br>56.5 % Male | 51.4 % Female,<br>48.6% Male | $X^2 (1, N = 162) = 1.01$ |
| SES Neighborhood Score | 9.54 (± 0.74) | 9.51 (± 0.74) | $t(159) = -0.21$ |
| Antidepressant Status | 15% | 9% | $X^2 (1, N = 162) = 1.62$ |
| WAB-AQ | 77.15 (± 22.38) | — | — |
| NIHSS Total Motor Score | 1.28 (± 1.93) | — | — |
| SIS Cognitive Score | 72.30 (± 19.07) | — | — |
| SIS Social Participation Score | 68.07 (± 25.01) | — | — |
| SIS Recovery Score | 68.04 (± 17.60) | — | — |
| SIS Physical Score | 72.78 (± 24.46) | — | — |
| BDI | 7.46 (± 6.25) | 4.03 (± 5.29) | — |

Abbreviations: SES, Socioeconomic Status; WAB-AQ, Western Aphasia Battery-Aphasia Quotient; NIHSS, National Institutes of Health Stroke Scale; SIS, Stroke Impact Scale; BDI, Beck Depression Inventory-II
\* $p < 0.05$ , \*\* $p < 0.01$ , \*\*\* $p < 0.001$

Years of education for each participant was calculated based on their highest degree earned, using a scoring scale by the National Institutes of Health (high school = 12, college = 16, graduate/professional degree = 18, JD = 19, MD = 20, PhD = 21). Socio-economic status was measured using the neighborhood score which was calculated based on the poverty levels associated with the current zip code. All participants provided current medication lists to determine whether they took antidepressant medications.

### Primary Measures of Interest

#### Beck Depression Inventory-II (BDI-II)

The Beck Depression Inventory-II (BDI-II) was administered to both controls and stroke survivors (Beck et al., 1996). The BDI-II is a 21-item self-report questionnaire that consists of 21 statements that measure the severity of depression. Participants select the statement that best describes how they have felt in the past two weeks, including the day of testing. Items are scored on a scale of 0 to 3 and total scores range from 0-63, with higher scores reflecting more severe depression (Beck et al., 1996). We omitted BDI-II items 16 (changes in sleeping patterns) and 18 (changes in appetite) from the current analyses, as previous literature has found this omission to better fit post-stroke and other neuropsychiatric populations (Lerdal et al., 2014). The control participants completed the questionnaire either on paper or on an iPad and were permitted to ask the tester if they had any questions. For stroke survivors, trained study personnel showed the questions to the stroke survivors, read them aloud, and explained each question and choices in case of reading comprehension difficulties.

#### Stroke Impact Scale 3.0 (SIS-3.0)

The 59-item Stroke Impact Scale 3.0 (SIS-3.0) was administered to stroke survivors (Duncan et al., 2003). The SIS-3.0 assesses stroke-related quality of life of stroke survivors (alternately framed as domain specific stroke-related disability) in eight domains: Strength, Hand Function, Activities of Daily Living (ADL) and Instrumental Activities of Daily Living (IADL), Mobility, Memory, Communication, Emotion, and Social Participation (Duncan et al., 2003). Multiple items contribute to each domain score. All items use a 5-point Likert scale and a total score is calculated from 0 to 100, where higher scores indicate better quality of life. There is also an item that measures perceived overall stroke recovery on a scale of 0 to 100. Subsequent research has demonstrated that a four domain structure exhibits better construct validity and reliability than the original eight domain structure (Gandolfi et al., 2021; Vellone et al., 2015). This new structure includes the following domains: Physical (combining Strength, Hand Function, ADL/IADL, and Mobility), Cognitive (combining Communication and Memory), Emotional (corresponding to the original Emotional domain), and Social Participation (corresponding to the original Social Participation domain). We included the Physical, Cognitive, Social Participation domains, and the Recovery score in our analyses, but omitted the Emotional domain, as the items are similar to the BDI-II because they are intended to measure similar constructs. The SIS 3.0 was administered by trained study personnel, who showed the questions to the stroke survivors, read them aloud, and provided a visual analog scale as an additional comprehension aid.

#### Measures of language and physical impairment

To directly measure language impairment without self-report, we administered the Western Aphasia Battery-Revised and used the Aphasia Quotient (WAB-AQ), which is often described as a measure of overall aphasia severity (Kertesz, 2022). To measure physical impairment without self-report, we administered the NIH Stroke Scale (NIHSS) and calculated a total motor score as the sum of the upper and lower extremity motor items.

### Statistical Analyses

Between-group *t*-tests and chi-squared tests were used to assess for differences in continuous and categorical demographic variables respectively between the groups. Spearman correlations were used to examine bivariate relationships between all continuous measures. Mann-Whitney U tests for race, gender, and medication status were used to examine differences in relation to BDI-II scores. An ordinal regression (Model 1) compared BDI-II total scores between stroke survivors and controls, with five demographic factors (age, education, gender, race, and socioeconomic status) and antidepressant medication status (yes/no) as binary factors or covariates as appropriate. A second ordinal regression (Model 2) tested BDI-II scores in stroke survivors as a function of the same demographic factors, plus SIS scores (4 subscales), time-since-stroke (in months), WAB-AQ, and NIHSS Total Motor score. Significance was set *a priori* at *p* < .05. All data was analyzed using SPSS Statistics 29.0.

## Results

The stroke and control groups were significantly different on education, but not on age, gender, race, SES, or antidepressant medication status (Table 1). The stroke group included a wide range of aphasia severity based on the WAB-AQ (range 13.7 to 100 out of 100), but was skewed toward mild deficits, with 41.3% of stroke survivors in the “mild aphasia” range on the WAB and 28.3% in the “no aphasia” range (>93.8). Notably, the WAB is known to lack sensitivity to mild aphasia (Silkes et al., 2021), and only 8.7% of stroke survivors reported no ongoing communication difficulties on the SIS. Scores on the BDI-II were relatively mild overall, with 94.3% of controls and 88.0% of stroke survivors in the range of “no depression or minimal depression,” 4.3% and 7.6% in the range of “mild depression,” and 1.4% and 4.3% in the range of “moderate depression” based on standard BDI-II cutoffs.

Bivariate Spearman correlations of the demographic measures with BDI-II revealed no relationships in the control group (Table 2). In the stroke group, bivariate Spearman correlations identified that higher BDI-II scores (greater depression symptoms) were associated with lower SIS Cognitive, Social Participation, and Recovery scores (p<.05, Bonferroni corrected for 49 comparisons, Table 3). At an uncorrected threshold, higher BDI-II scores were also associated with lower WAB-AQ, younger age, and lower education. The SIS Physical score and the NIHSS Total Motor score were unrelated to BDI-II scores. Correlations also revealed expected relationships among the disability and impairment measures (Table 3).

**Table 2.**
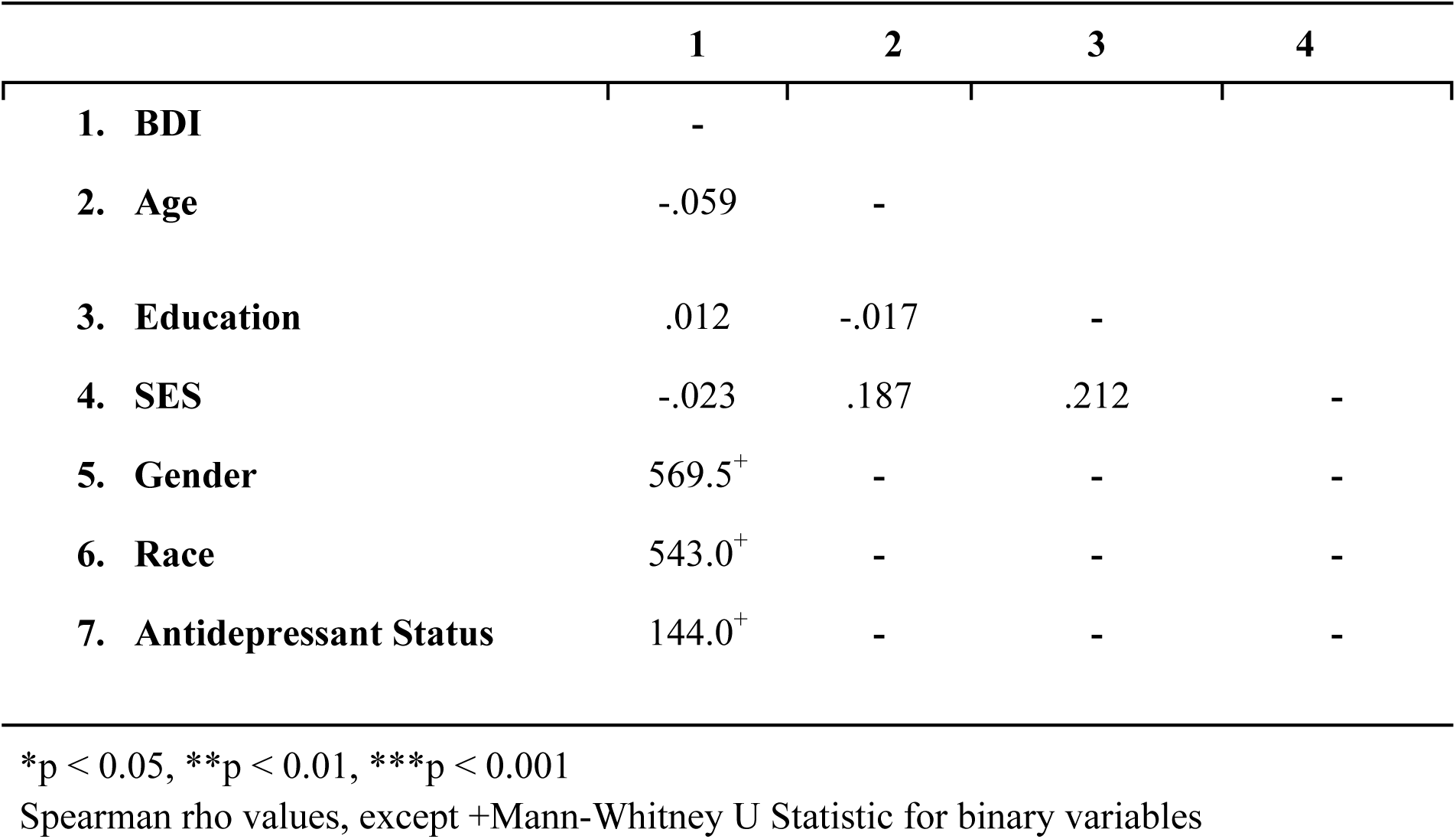
Relationships among variables of Interest (Controls)

|  | **1** | **2** | **3** | **4** |
| --- | --- | --- | --- | --- |
| **1. BDI** | - |  |  |  |
| **2. Age** | -.059 | - |  |  |
| **3. Education** | .012 | -.017 | - |  |
| **4. SES** | -.023 | .187 | .212 | - |
| **5. Gender** | 569.5<sup>+</sup> | - | - | - |
| **6. Race** | 543.0<sup>+</sup> | - | - | - |
| **7. Antidepressant Status** | 144.0<sup>+</sup> | - | - | - |

\*p < 0.05, \*\*p < 0.01, \*\*\*p < 0.001
Spearman rho values, except +Mann-Whitney U Statistic for binary variables

**Table 3.**
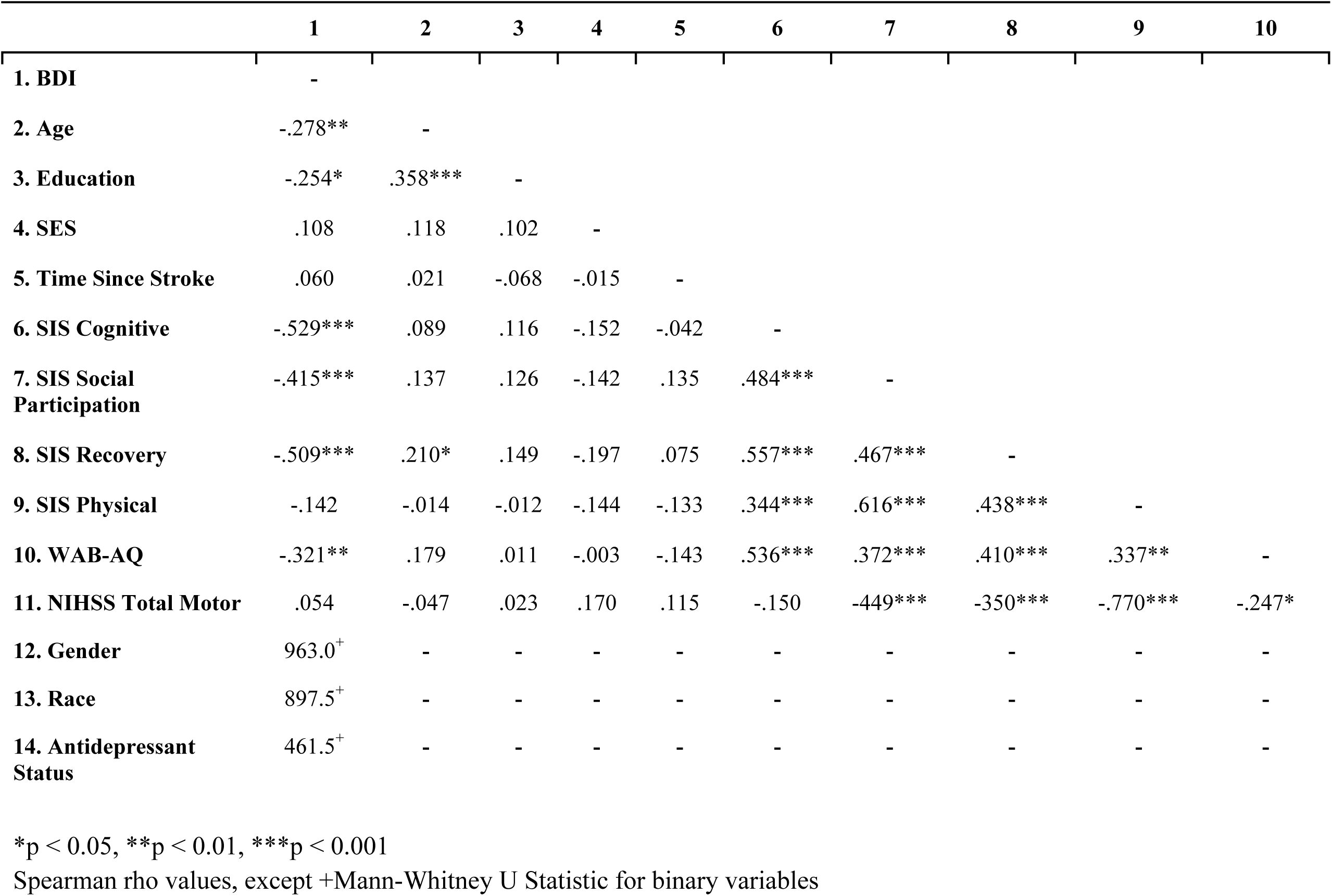
Relationships among variables of Interest (Stroke)

|  | 1 | 2 | 3 | 4 | 5 | 6 | 7 | 8 | 9 | 10 |
| --- | --- | --- | --- | --- | --- | --- | --- | --- | --- | --- |
| **1. BDI** | - |  |  |  |  |  |  |  |  |  |
| **2. Age** | -.278** | - |  |  |  |  |  |  |  |  |
| **3. Education** | -.254* | .358*** | - |  |  |  |  |  |  |  |
| **4. SES** | .108 | .118 | .102 | - |  |  |  |  |  |  |
| **5. Time Since Stroke** | .060 | .021 | -.068 | -.015 | - |  |  |  |  |  |
| **6. SIS Cognitive** | -.529*** | .089 | .116 | -.152 | -.042 | - |  |  |  |  |
| **7. SIS Social Participation** | -.415*** | .137 | .126 | -.142 | .135 | .484*** | - |  |  |  |
| **8. SIS Recovery** | -.509*** | .210* | .149 | -.197 | .075 | .557*** | .467*** | - |  |  |
| **9. SIS Physical** | -.142 | -.014 | -.012 | -.144 | -.133 | .344*** | .616*** | .438*** | - |  |
| **10. WAB-AQ** | -.321** | .179 | .011 | -.003 | -.143 | .536*** | .372*** | .410*** | .337** | - |
| **11. NIHSS Total Motor** | .054 | -.047 | .023 | .170 | .115 | -.150 | -.449*** | -.350*** | -.770*** | -.247* |
| **12. Gender** | 963.0<sup>+</sup> | - | - | - | - | - | - | - | - | - |
| **13. Race** | 897.5<sup>+</sup> | - | - | - | - | - | - | - | - | - |
| **14. Antidepressant Status** | 461.5<sup>+</sup> | - | - | - | - | - | - | - | - | - |

\*p < 0.05, \*\*p < 0.01, \*\*\*p < 0.001
Spearman rho values, except +Mann-Whitney U Statistic for binary variables

**Table 4.** Model 1 Statistics (stroke vs. control)

|  | Estimate | Wald | 95% CI |
| --- | --- | --- | --- |
| Group (Stroke vs. Control) | $\beta = -1.232^{***}$ | 17.087 | (-1.817, -0.648) |
| Age | $\beta = -0.027^*$ | 4.678 | (-0.052, -0.003) |
| Education | $\beta = -0.104$ | 3.678 | (-0.211, 0.002) |
| Race | $\beta = -0.382$ | 1.453 | (-1.003, 0.239) |
| Gender | $\beta = -0.269$ | 0.893 | (-0.827, 0.289) |
| SES Neighborhood Score | $\beta = 0.305$ | 2.194 | (-0.099, 0.709) |
| Antidepressant Status | $\beta = 0.036$ | 0.007 | (-0.792, 0.864) |
| Overall model fit | $X^2 = 31.44^{***}$ | — | — |

\* $p < 0.05$ , \*\* $p < 0.01$ , \*\*\* $p < 0.001$

**Table 5.** Model 2 Statistics (stroke only)

|  | Estimate | Wald | 95% CI |
| --- | --- | --- | --- |
| Number of Participants | — |  |  |
| Age | $\beta = -0.028$ | 2.223 | (-0.064, 0.009) |
| Education | $\beta = -0.093$ | 1.611 | (-0.236, 0.051) |
| Race | $\beta = -0.424$ | 0.936 | (-1.282, 0.435) |
| Gender | $\beta = 0.298$ | 0.522 | (-0.510, 1.107) |
| SES Neighborhood Score | $\beta = -0.050$ | 0.032 | (-0.601, 0.500) |
| Time since Stroke (months) | $\beta = 0.011^{**}$ | 6.165 | (0.002, 0.019) |
| Antidepressant Status | $\beta = 0.697$ | 1.668 | (-0.361, 1.755) |
| WAB-AQ | $\beta = -0.002$ | 0.027 | (-0.022, 0.019) |
| NIHSS Total Motor | $\beta = 0.017$ | 0.012 | (-0.286, 0.320) |
| SIS Cognitive | $\beta = -0.034^{**}$ | 5.890 | (-0.061, -0.006) |
| SIS Social Participation | $\beta = -0.038^{***}$ | 11.401 | (-0.061, -0.016) |
| SIS Recovery | $\beta = -0.054^{***}$ | 12.295 | (-0.085, -0.024) |
| SIS Physical | $\beta = 0.036^{**}$ | 6.013 | (0.007, 0.065) |
| BDI | — | — | — |
| Overall model fit | $X^2 = 63.60^{***}$ | — | — |

\* $p < 0.05$ , \*\* $p < 0.01$ , \*\*\* $p < 0.001$

Model 1 used an ordinal regression to test for differences in depressive symptoms between stroke survivors and controls with demographic factors and antidepressant medication status as covariates. As expected, Model 1 demonstrated that stroke survivors had higher BDI-II scores than controls (β = −1.232, p < 0.001). Younger age (β = −0.027, p < 0.05) was also associated with higher BDI-II scores. Education was not a significant predictor, but trended in the direction of lower education relating to higher BDI-II scores (β = −0.104, p = 0.055).

Model 2 used an ordinal regression to test BDI-II scores in stroke survivors as a function of the same demographic factors, plus the four SIS domain scores (Physical, Cognitive, Social Participation, Recovery), time since stroke, language impairment (WAB-AQ), and motor impairment (NIHSS Total Motor score). In Model 2, lower scores on the SIS Cognitive (β = - 0.034, p < 0.01), Social Participation (β = −0.038, p < 0.001) and Recovery (β = −0.054, p < 0.001) scores were associated with higher depression. Although higher scores on the SIS physical (β = 0.036, p < 0.01) were associated with higher depression, this likely resulted from covariance among predictors, as it was unrelated to depression scores in a bivariate spearman correlation (*r*_s_ (92) = −0.142, *p* = 0.176), with a negative coefficient (the opposite direction compared to the regression beta value). Greater time since stroke related to higher depression symptoms in the model (β = 0.011, p < 0.013), but this result should also be interpreted with caution, as time since stroke was unrelated to BDI-II scores in a bivariate spearman correlation (*r*_s_ (92) = 0.060, *p* = 0.570). To confirm the robustness of the overall Model 2 results, we repeated the model leaving out the SIS Physical score due to the likely spurious relationship. Lower SIS Cognitive, Social Participation, and Recovery scores were significantly related to higher BDI-II scores as in the full model, but time since stroke was no longer a significant predictor.

## Discussion

Since the experience of living with stroke-related disability differs depending on the types of deficits caused by the stroke, research on PSD may benefit from examining specific populations with common stroke locations or deficit patterns. To our knowledge, this is the first study to examine the relationship between symptoms of depression and multiple domains of disability, impairment, and demographic factors specifically in a left-hemisphere stroke population with a history of aphasia. Previous studies have not examined the SIS-3.0, a comprehensive measure of stroke-related quality of life and disability, in relation to depression. Using the SIS-3.0 to assess multiple dimensions of disability after stroke provides a broad view of potential correlates of depression. The results indicate that the primary drivers of depression in chronic left-hemisphere stroke survivors include self-perceived disability in communication and cognition, low social participation, and self-perceived poor recovery.

As expected, left-hemisphere stroke survivors reported greater depression symptoms than the controls, consistent with prior studies showing high rates of depression after stroke (Liu et al., 2023). The raw difference in BDI-II scores between groups was 3.4 points, but after accounting for demographic factors and antidepressant medication status, the estimated difference reduced to 1.2 points, a small but highly significant difference. The finding that depression symptoms remain greater than controls in the chronic period warrants further research on depression in this phase of recovery, along with longitudinal research to determine how depression symptoms and the factors driving them may change over time.

The model including both groups and bivariate correlations in the stroke group identified a relationship between age and depression symptoms, consistent with prior studies examining emotional wellbeing across the lifespan that have found higher age to be associated with lower depression (McCarthy et al., 2016). Some theories, such as the Socioemotional Selectivity Theory (Carstensen, 2006), suggest that older individuals prioritize more meaningful day-to-day activities, relationships, and events. Older adults also may have life experience facilitating higher emotion regulation, less fixation on negative emotion, and more positive social judgment (Charles & Carstensen, 2010). Alternatively, older individuals may under-report depressive symptoms due to stigma around mental health or clinicians and older adults normalizing the trajectory of increased depressive symptoms with age (Lutz & Van Orden, 2020; Lyness et al., 1995). Regardless, neither age nor other demographic variables were significant predictors of depression symptoms in stroke survivors when considered alongside stroke-related disability and impairment. This is likely because stroke-related disabilities are the primary correlates of depression symptoms and overwhelm the variance associated with age. As such, the observed association between depression symptoms and age is not specific to PSD, and likely reflects factors associated with lower depression symptoms in an aging population more generally.

In the second model, we found that lower scores on the SIS Cognitive, Social Participation, and Recovery scores (indicating more cognitive or communication disability, lower social participation, and less perceived recovery) were associated with higher depression scores. These measures were also the only variables related to depression symptoms in bivariate correlations after correction for multiple comparisons, reinforcing their robust relationship with depression symptoms in this sample. In contrast, physical disability was unrelated to depression in bivariate correlations, and spuriously anticorrelated with depression when included in a model with other predictors. These findings suggest that in a chronic stroke population recovering from aphasia, physical disability may not be a primary driver of depression even though the motor impairment caused by left-hemisphere stroke most often affects the dominant arm. This finding contrasts with other studies of PSD that found that measures of physical disability or motor impairment related to depression (e.g., Choi et al., 2023; De Ryck et al., 2014). Prior studies have not separated participants based on the hemisphere affected by their stroke, resulting in a mixture of different types of non-motor difficulties faced by participants that could bias findings toward common issues faced by the group, such as physical disability. Prior studies have also most often assessed depression within the first few weeks or months after stroke, in contrast to our sample of chronic stroke survivors who may have accepted their physical limitations over time. In addition, many prior studies excluded people with significant aphasia, precluding them from identifying communication disability or speech and language impairment as important correlates of depression symptoms. Alternatively, perhaps our sample was biased because the overarching study in which the current data were collected focused on aphasia. As such, individuals who participated may naturally focus primarily on their communication and cognitive disability rather than physical limitations. Thus, the current results should not be taken to indicate that physical disability is unrelated to depression in a general stroke population. Further research should address this question in other research contexts to confirm our results.

In our sample, depression symptoms related to disability in communication and cognition as measured by the SIS Cognitive score. Of note, the SIS Cognitive score includes items that were originally grouped into separate Communication and Memory domains. However, several of the items in the Memory domain may be affected by communication difficulties (e.g., “In the past week, how difficult was it for you to remember things that people just told you?”). Prior factor analyses have demonstrated that the Communication and Memory domain items measure one underlying factor (Vellone et al., 2015), resulting in the combined Cognitive domain score used here. Therefore, it is not possible to determine whether communication versus memory or other cognitive limitations drove the relationship between the Cognitive score and the BDI-II. However, given that aphasia is the primary cognitive syndrome associated with left hemisphere strokes, and the majority of items composing this score relate at least in part to communication ability (11 out of 14 items), it is likely that communication disability was the primary driver of this relationship.

Notably, language impairment as measured by the WAB-AQ did not relate to depression symptoms, except in a bivariate correlation that did not survive multiple comparisons correction. The SIS is a subjective measure which probes self-reported activities from daily life, whereas the WAB, a more objective measure of speech and language impairment, is scored by a clinician or a researcher in a controlled testing environment. Intuitively, an individual’s own perception of their ability to communicate in daily life will correspond more closely to symptoms of depression such as low mood, worthlessness, and feelings of self-criticalness than performance on tests intended to assess processing impairment devoid of real-world communication contexts (e.g., naming as many animals as possible in one minute or following unintuitive sequences of commands, such as “with the book, point to the comb.” Some previous literature examining associations between chronic post-stroke aphasia, communication, and depression have found functional communication (communication relevant to daily life) measured by either subjective and objective scales to predict depressive symptoms (Ashaie et al., 2019; Døli et al., 2017), while others found that it did not predict PSD (Bueno-Guerra, 2023; Pompon et al., 2022). Studies that have focused on this relationship are somewhat difficult to compare given differences in culture, location, stroke chronicity, and measures. However, we know that the inability to communicate due to aphasia negatively impacts quality of life (Grönberg et al., 2024; Spaccavento et al., 2013) and lower quality of life predicts increased depressive symptoms (Herman et al., 2002). Specifically, the inability to communicate frequently results in loss of autonomy, inability to return to work, and social isolation or lack of belonging within a community (Burfein et al., 2024; Dalemans et al., 2010; Palstam et al., 2019). Individuals with aphasia report difficulty accessing mental health services, including individual psychotherapists and group interventions, that can meet their needs because of communication disability (Worrall et al., 2016). Thus, self-perceived communication ability may be more relevant to daily life functioning and participation and may better predict depressive outcomes as compared to a test like the WAB, which may not be as sensitive to these real-world challenges.

The SIS Social Participation score, a measure of social relationships and life participation (e.g., how much have you been limited in work, social activities, ability to help others, role as a friend?), also related to depression symptoms. Importantly, the SIS Social Participation and Cognition scores were both significantly related to depression symptoms even when included in the same model. This demonstrates that their relationships with depression are at least partly independent. There are many potential causes of decreased social participation after stroke, including not just communication and cognitive impairment, but also limitations in mobility, lack of support, and a fear of judgment of impairment from self and others (Della Vecchia et al., 2021; Desrosiers et al., 2006; Wallace, 2010). Decreased social participation is also associated with higher risk and presence of depressive symptoms in adults outside the context of stroke (Holtfreter et al., 2017).

The SIS Recovery factor assesses how the individual perceives their overall recovery after stroke. Poor self-perceived recovery could relate to either a real lack of improvement in abilities over time or unrealistic expectations for recovery. Either might be expected to affect an individual’s mental well being, perhaps eliciting feelings of hopelessness or helplessness and a lack of motivation given prior experiences of recovery not meeting expectations. Alternatively, a lack of confidence or low self-perception related to depression occurring for other reasons might lead to perceptions that one’s recovery is inadequate.

The theorized bidirectional relationship between depression symptoms and self-reports of recovery described above applies more broadly to other self-reported disability measures as well. While we have focused above on the ways in which disability might cause depression symptoms, the opposite causal relationship is also likely at play to some degree. For example, evidence suggests a bidirectional relationship between depression and social isolation, in that low social participation can cause depression and depression can reduce social participation (Ding et al., 2022). Since the SIS is a self-reported measure, greater depression might also lead to poorer self-perception with regard to abilities, and thus over-reporting disability. We believe the primary direction of effect in this study is that disability drives depression because the SIS Physical score did not relate to depression, and each of the other SIS scores contributed independent variance to depression scores. If the primary direction of effect was that depression resulted in greater self-perceived disability, one might expect this to be reflected uniformly across domains rather than the differential pattern observed here. Future studies should assess the longitudinal trajectories of self-perceived disability and depression symptoms over the course of stroke recovery, which may help to disentangle the causal direction of the relationships. For example, if depression symptoms precede a reduction in self-rated disability, this would suggest that depression drives low disability scores. Interventions to improve depression symptoms may also be implemented to examine whether alleviation of depression symptoms results in less perceived disability.

One notable limitation of this study is a potential sampling bias, as participants who were severely depressed would most likely not volunteer to participate in research. This may be reflected by the relatively mild symptoms of depression in the sample overall. The SIS-3.0 and BDI-II were administered by study personnel for the stroke participants whereas the controls completed the BDI-II unmonitored. This difference may allow for stroke participants to gain a more clear understanding of the questions, as they are also provided with pictures to help with comprehension, and spend more time thinking through responses. This difference could potentially result in a difference in scores between the groups. While participants included a relatively large proportion of Black participants, a minoritized group who experience well-documented disparities in stroke care and stroke outcomes, the focus of the overarching project on aphasia precluded participation by members of minority groups who are not native English speakers.

Overall, these findings highlight important disability factors related to depression after left-hemisphere stroke causing aphasia, which may differ from the factors contributing to depression in individuals without history of aphasia. Individuals with aphasia form a plurality of stroke survivors and should not be systematically excluded from studies of depression; rather, communication support should be given to facilitate participation by all individuals except those with the most severe comprehension deficits. Along with being excluded from research on depression, individuals with aphasia are often excluded from participation in psychotherapy due to misconceptions that they will not be able to participate because of communication barriers. Specific training programs on aphasia should be developed for mental healthcare providers to facilitate their ability to provide care for individuals with aphasia and depression. Our findings demonstrate that there is a need for such care even long after stroke and suggest potential drivers of depression that could be targeted in psychotherapy, for example aiming to increase acceptance of disabilities and a changed reality through Acceptance and Commitment Therapy (Large et al., 2020), or working on techniques to increase emotion regulation, mood, and life participation through Cognitive Behavioral Therapy (Kneebone, 2016). These treatments have been adapted for individuals with brain injury, particularly focused on cognitive and communication disabilities, but it is critical for mental healthcare providers to receive more training in working with these populations.

## Data Availability

Data will be made available upon reasonable request.

## Non-standard Abbreviations and Acronyms

BDI-II: Beck Depression Inventory-II
SIS: Stroke Impact Scale 3.0
WAB-AQ: Western Aphasia Battery Aphasia Quotient
NIHSS: National Institutes of Health Stroke Scale
SES: Socioeconomic Status

## Acknowledgements

Thank you to our participants and non-author data collectors: Alycia Laks, Trini Kelly, Sarah Snider, Candace van der Stelt, Elizabeth Lacey, and Elizabeth Dvorak.

